# Overlooked Trustworthiness of Explainability in Medical AI

**DOI:** 10.1101/2021.12.23.21268289

**Authors:** Jiajin Zhang, Hanqing Chao, Mannudeep K. Kalra, Ge Wang, Pingkun Yan

## Abstract

While various methods have been proposed to explain AI models, the trustworthiness of the generated explanation received little examination. This paper reveals that such explanations could be vulnerable to subtle perturbations on the input and generate misleading results. On the public CheXpert dataset, we demonstrate that specially designed adversarial perturbations can easily tamper saliency maps towards the desired explanations while preserving the original model predictions. AI researchers, practitioners, and authoritative agencies in the medical domain should use caution when explaining AI models because such an explanation could be irrelevant, misleading, and even adversarially manipulated without changing the model output. AI researchers, practitioners, and authoritative agencies in the medical domain should use caution when explaining AI models because such an explanation could be irrelevant, misleading, and even adversarially manipulated without changing the model output.

## I Introduction

Explainability is a pillar in supporting applications of artificial intelligence and machine learning (AI/ML) in medicine^1^. Understanding the how and why AI models make particular decisions is critical for building trust in any AI-driven application^2^. There is a large body of research papers on methods revealing how AI works^3,4^. The Food and Drug Administration (FDA) is the federal agency responsible for clearing medical AI devices for clinical use in the United States. FDA stresses trustworthiness as a key factor for adopting and evolving medical AI/ML devices, a viewpoint endorsed by more than 40 other countries^5^. While the research community appreciates the development of explainable AI (XAI), we often focus on how to explain an AI model but have not answered an important question - can the explainability be trusted? The trustworthiness of AI-based saliency methods has been overlooked. This has left potential risks to AI-based applications in the medical field.

To understand the extent of these concerns, here we demonstrate worrisome phenomena with saliency maps in disease diagnosis from medical images through both case studies and statistical analysis. Saliency maps, also commonly referred to as heat maps, are the most commonly used method for AI explainability^6^. The research community is increasingly aware that the output of an AI model could abruptly change with subtle perturbations to the input, especially adversarial attacks^7^. Little research has been done to understand the impact of tampered inputs on the explainability of AI^8^. It is therefore imperative to pay attention to the trustworthiness of explainability in medical AI.

## II Methods

### 2.1 Saliency Maps Can Be Misleading

We compiled a representative list of medical AI papers utilizing the various saliency map generation methods (referred to as saliency methods for conciseness) for their explainability (https://github.com/DIAL-RPI/Trustworthiness-of-Medical-XAI). In this list, seven most commonly used saliency methods, including Vanilla Grad^9^, Grad x Image^10^, GradCAM^6^, Guided-GradCAM^6^, IG^11^, SG^12^ and XRAI^13^, were selected with applications in medical AI applications. We used a public chest radiograph dataset, CheXpert,^14^ and a widely used baseline model, DenseNet^15^, to demonstrate potential problems with saliency maps. Technical details of model training are publicly available at (https://github.com/DIAL-RPI/Trustworthiness-of-Medical-XAI). We evaluated the model performance by the area under the receiver operating characteristic curve (AUC). Following the original work of CheXpert^14^, we focused on five observations: (a) Atelectasis, (b) Cardiomegaly, (c) Consolidation, (d) Edema, and (e) Pleural Effusion. Our DenseNet achieved a mean AUC of 0.895, slightly surpassing the performance presented in the original paper^14^.

The objective of our study is to demonstrate that saliency maps of an AI model can be easily manipulated by applying human imperceptible perturbations to an input image without noticeable change of the model output. **Fig. 1** illustrates our study. Given an original input image and the trained AI model, an adversarial image can be generated by adding human-imperceptible perturbations to the original image. More specifically, the adversarial counterpart can be computed by minimizing the loss

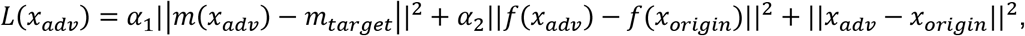

where *m*(*) represents a certain saliency method, *f*(*) denotes the AI model, the hyperparameter *α*_1_ and *α*_2_ were empirically set to 10^14^ and 10^3^, respectively, in our experiment.

**Figure 1.**
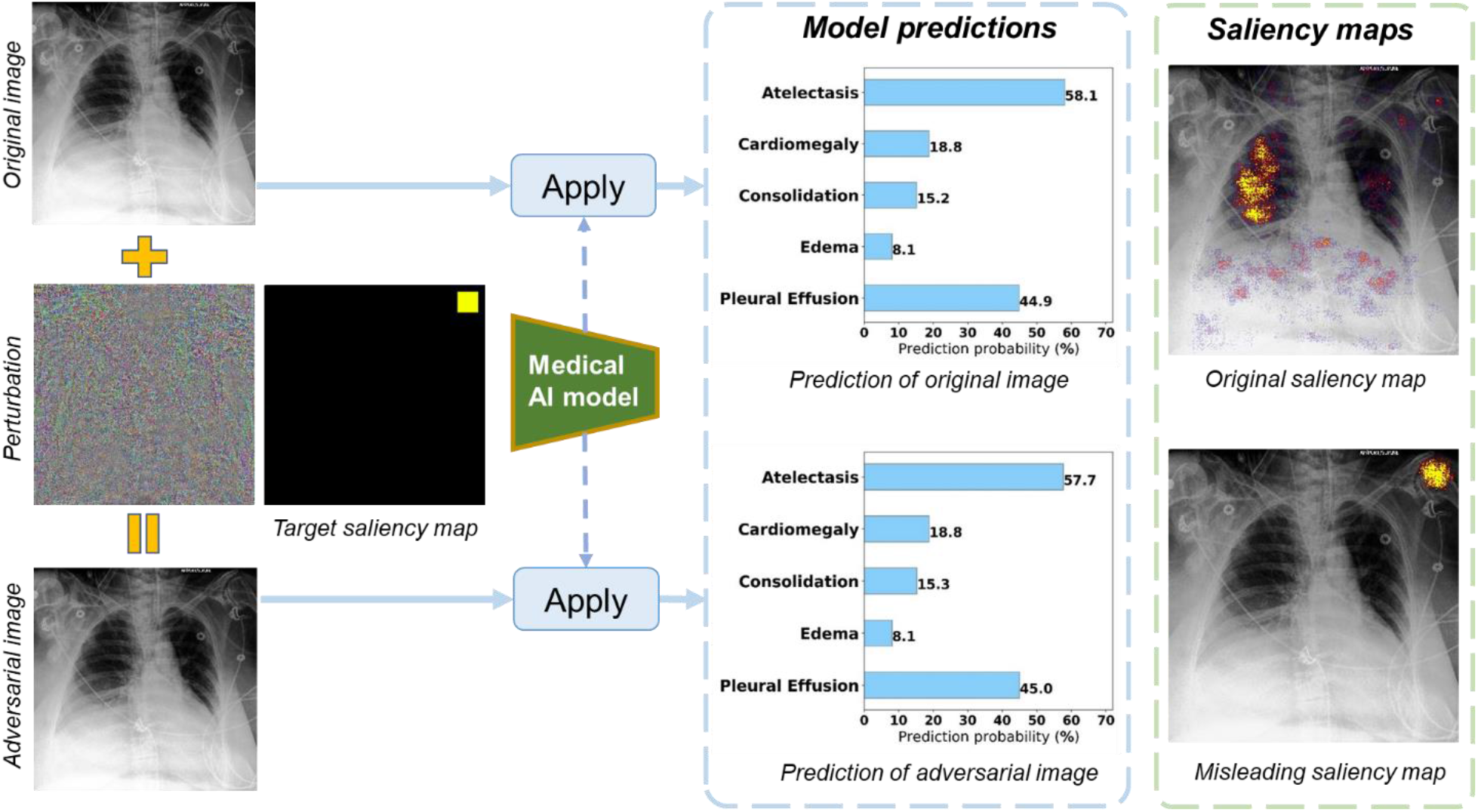
*Our study illustrated by an example, where an adversarial image with human-imperceptible tampers the saliency map without changing the model predictions. The data and model used in this example are available at* (https://github.com/DIAL-RPI/Trustworthiness-of-Medical-XAI).

By minimizing the loss through manipulating an adversarial image *x*_*adv*_, we can have the associated saliency map *m*(*x*_*adv*_) very close to an arbitrarily designed target saliency map *m*_*target*_. At the same time the adversarial image and the model output only deviate slightly from the original image and the corresponding output, respectively. As shown in Fig. 1, when the original image and its adversarial counterpart are fed into the same medical AI model, the outputs of the AI model are in excellent agreement. However, the saliency map of the adversarial image has been tampered towards the intentionally designed pattern. In this case, we specifically made the saliency region irrelevant to the model prediction, which is located on the top right corner of the image. This finding is concerning as it shows not only the saliency map can be completely irrelevant but also such a map can be purposely manipulated to a desired target region. This safety issue significantly reduces the trustworthiness of the explainability of saliency maps.

### 2.2 The Concern is Universal

Next, we underline that this concern is not only universal to the commonly used saliency methods summarized above but also the adversarial images are transferrable across different saliency methods. Specifically, we demonstrate the misleading effects on all the seven representative saliency methods in **Fig. 2**. The first row of **Fig. 2** shows the saliency maps generated on the original input image. The highlighted regions correspond to the observation of atelectasis annotated for this frontal view X-ray image. The adversarial images can be generated for each method following our method described above, which are publicly available at (https://github.com/DIAL-RPI/Trustworthiness-of-Medical-XAI/samples). These adversarial images did not change the model predictions but did falsify saliency maps consistently as shown in the third row of **Fig. 2**. For all the seven methods, the adversarial images succeeded in shifting the saliency areas to the targeted top right corner of the image. In other words, even if multiple explanation methods produce consistent results, they may still be misleading.

**Figure 2.**
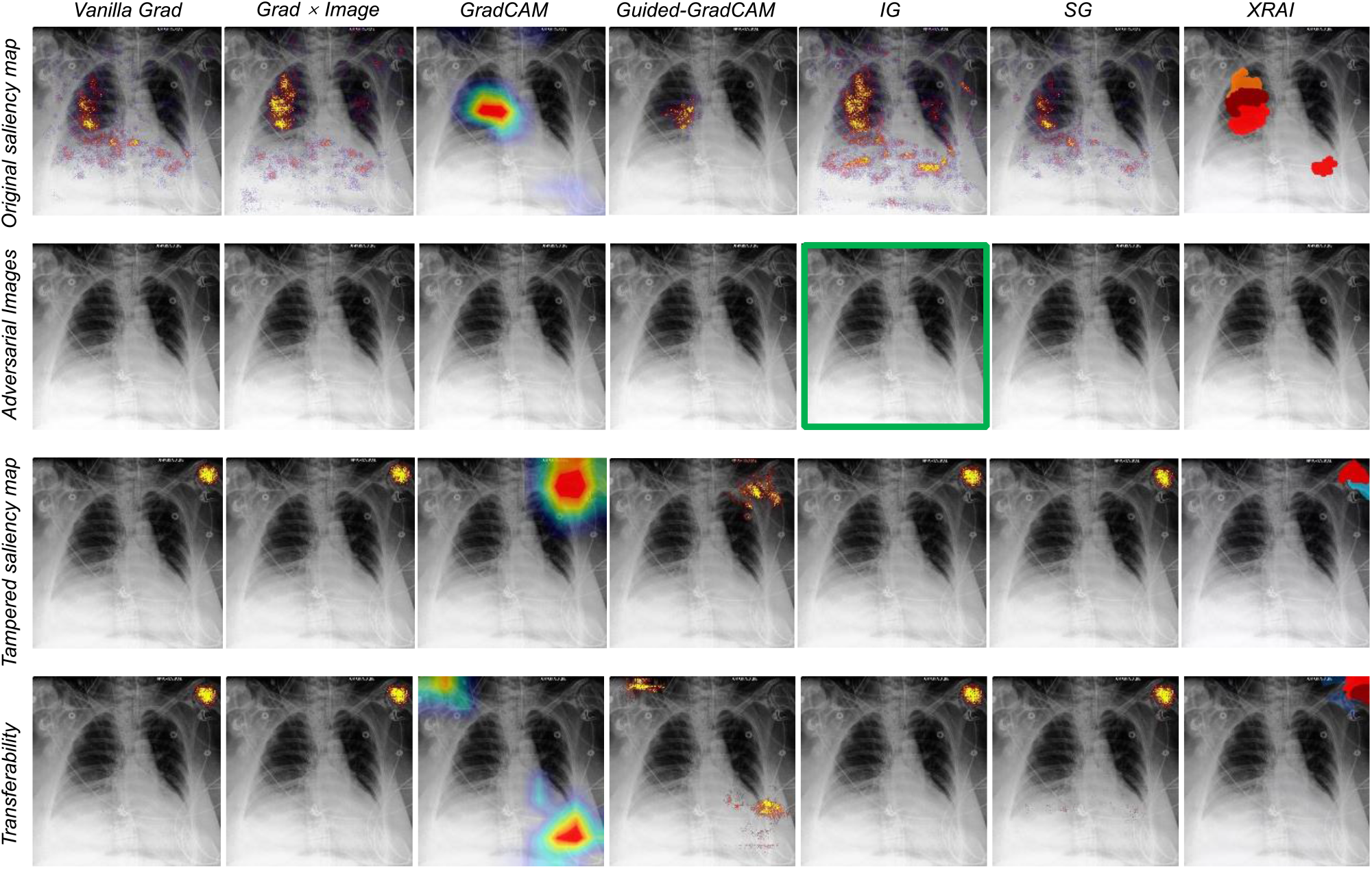
Collusion of explanation methods. **First row**: saliency maps generated using different methods on the same original input image; **Second row**: generated adversarial images for each saliency visualization method. **Third row**: tampered saliency maps using the above adversarial images; **Last row**: saliency maps generated on the adversarial image highlighted by the green box in the second row.

We further examined the existence of universal adversarial images by evaluating the transferability of the adversarial images across different XAI methods. We used the adversarial image generated for attacking IG as input and produced saliency maps using other methods. The last row of **Fig. 3** shows that the adversarial image towards IG can be well transferred to attack the other methods by shifting the attention areas. For majority of the methods, the attention area was moved to the targeted top-right corner of the image. More cases studies can be found at (https://github.com/DIAL-RPI/Trustworthiness-of-Medical-XAI).

**Figure 3.**
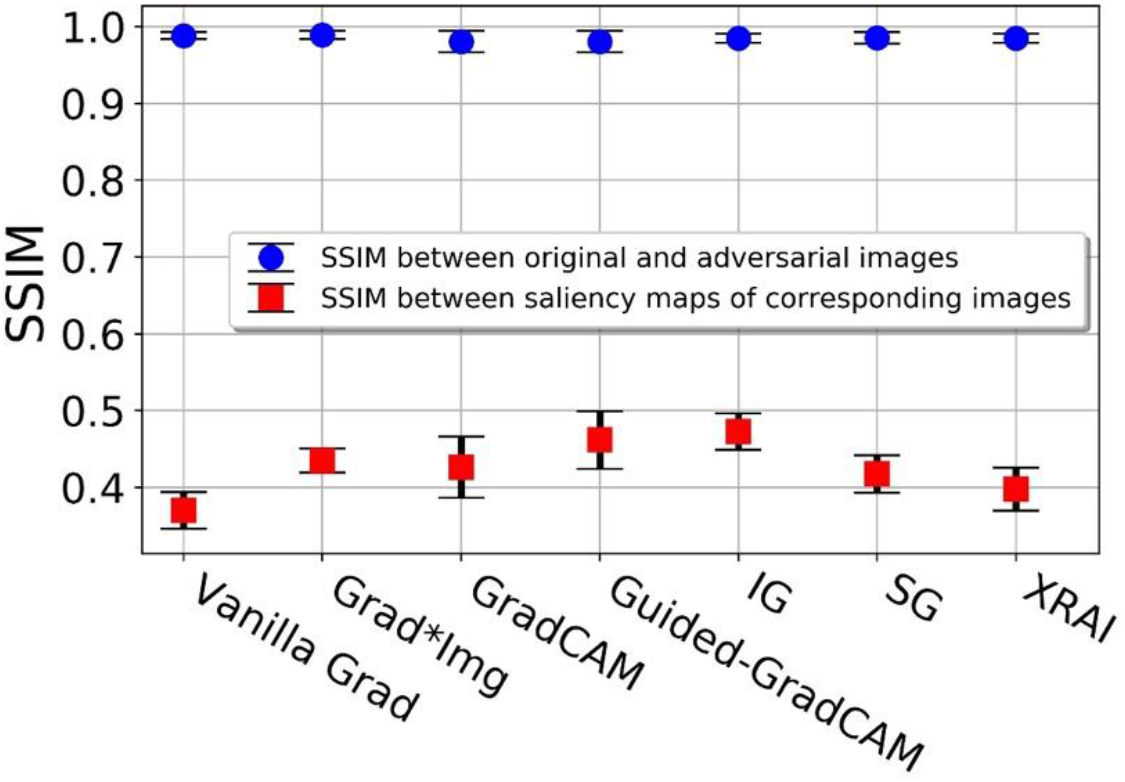
Comparation between original and adversarial results. Blue box plots: similarities between the original images and the adversarial images; and red box plots: similarities between the saliency maps for the original and adversarial inputs. The dots and squares in the plots denote the mean values, and the bars indicate the standard deviations.

### 2.3 Ensuring AI Model Prediction Consistence is Not Enough

We quantitatively evaluated the image similarities and saliency map similarities as well as the model prediction consistence on 200 chest radiographs from the CheXpert validation set. The similarities were assessed in terms of the structural similarity index measure (SSIM)^16^. The results in **Fig. 3** indicate that the tampered saliency maps are vastly different (mean SSIM<0.50) from the corresponding maps for the original images, while the adversarial images are remarkably close to the original images (mean SSIM>0.98). This indicates that the perturbations added to the adversarial images may be hardly noticeable by humans but they could cause significant changes to the saliency maps. Additional evaluation metrics including Mean-Square Error (MSE) and Pearson Correlation Coefficient (PCC) were also used to provide the same conclusion. The results are available at (https://github.com/DIAL-RPI/Trustworthiness-of-Medical-XAI).

In addition, we evaluated the effect of the adversarial images on the model performance measured by AUC, prediction consistency assessed by AUC change percentage relative to the original value of 0.895, and unsuccessful attack ratio. An attack is considered as failed if the SSIM between the tampered saliency map and the target map is below 0.6. The results in **Table 1** show that the AI model performed consistently under the original input images and adversarial images, with the performance change less than 2.5%. At the same time, the saliency maps were successfully manipulated towards the target with the unsuccessful attack rates all below 10% for all the XAI methods. These results demonstrate that model saliency maps as an explainability approaches can be purposely manipulated even the predictions remain consistent. That is, the correctness of the model predictions may not support the validity of explanations to the model.

**Table 1.**
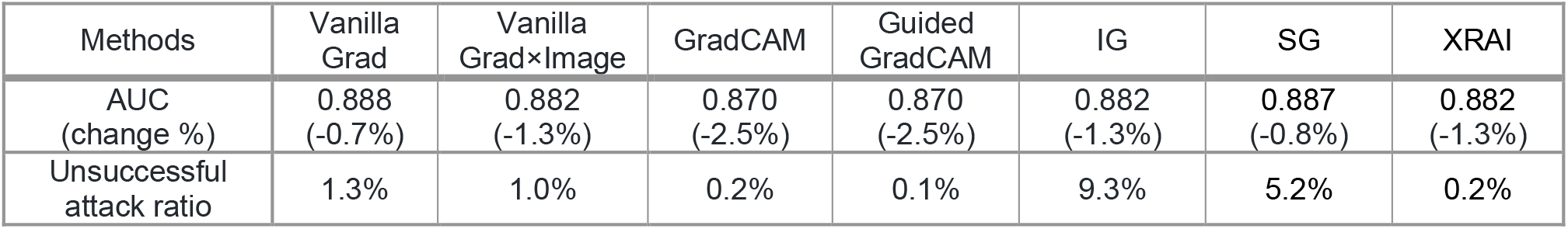
Saliency maps are successfully manipulated with only slight influence to the model performance.

| Methods | Vanilla Grad | Vanilla GradxImage | GradCAM | Guided GradCAM | IG | SG | XRAI |
| --- | --- | --- | --- | --- | --- | --- | --- |
| AUC (change %) | 0.888 (-0.7%) | 0.882 (-1.3%) | 0.870 (-2.5%) | 0.870 (-2.5%) | 0.882 (-1.3%) | 0.887 (-0.8%) | 0.882 (-1.3%) |
| Unsuccessful attack ratio | 1.3% | 1.0% | 0.2% | 0.1% | 9.3% | 5.2% | 0.2% |

## III Discussions and Conclusions

The input images in our experiment were adversarially perturbed to reveal severe safety issues of the existing XAI methods. Such perturbations could appear naturally in medical imaging due to numerous variations in commercial vendors, acquisition parameters, image reconstruction algorithms, patient characteristics, and so on. Also, malicious human hackers in the healthcare domain were reported before^17^, and their persistent existence should be a valid assumption. Given the limited evaluation of findings with most FDA cleared AI models, the reported problems could arise in presence of distracting or mimicking findings to the intended target for AI algorithms. Therefore, the findings in this paper suggest that AI researchers, practitioners and authoritative agencies in the medical domain must use caution when explaining AI models in their applications with saliency maps, and perhaps other types of explanation methods as well. Solid quality-assurance measures are urgently needed to increase and validate the trustworthiness of AI explainability, especially in the medical field.

## Data Availability

All data produced are available online at

https://stanfordmlgroup.github.io/competitions/chexpert/

